# Cost-Sensitive Machine Learning Classification for Mass Tuberculosis Screening

**DOI:** 10.1101/19000190

**Authors:** Ali Akbar Septiandri, Aditiawarman, Roy Tjiong, Erlina Burhan, Anuraj H. Shankar

## Abstract

Active screening for Tuberculosis (TB) is needed to optimize detection and treatment. However, current algorithms for verbal screening perform poorly, causing misclassification that leads to missed cases and unnecessary and costly laboratory tests for false positives. We investigated the role of machine learning to improve the predefined one-size-fits-all algorithm used for scoring the verbal screening questionnaire. We present a cost-sensitive machine learning classification for mass tuberculosis screening. We compared score-based classification defined by clinicians to machine learning classification such as SVM-RBF, logistic regression, and XGBoost. We restricted our analyses to data from adults, the population most affected by TB, and investigated the difference between untuned and unweighted classifiers to the cost-sensitive ones. Predictions were compared with the corresponding GeneXpert MTB/Rif results. After adjusting the weight of the positive class to 40 for XGBoost, we achieved 96.64% sensitivity and 35.06% specificity. As such, sensitivity of our identifier increased by 1.26% while specificity increased by 13.19% in absolute value compared to the traditional score-based method defined by our clinicians. Our approach further demonstrated that only 2000 data points were sufficient to enable the model to converge. Our results indicate that even with limited data we can actually devise a better method to identify TB suspects from verbal screening. This approach may be a stepping stone towards more effective TB case identification, especially in primary health centres, and foster better detection and control of TB.

## I. Introduction

Automated tuberculosis (TB) detection has specifically been studied for nearly two decades. One of such is by using rule-based approach from both clinical and laboratory criteria [1]. In a more recent approach, [2] were able to achieve 99.14% accuracy, 87.00% sensitivity, and 86.12% specificity in TB diagnosis using Artificial Immune Recognition System (AIRS) learned from supporting lab results. It is also shown in [3] that using artificial neural networks (ANN) could help them to reach more 94.5% sensitivity and 91.0% specificity in diagnosing pleural TB based on anamnesis variables and HIV status. Although a real-time screening alert has been done in [4], machine learning was not employed in that study. Therefore, we proposed to do so in this paper aiming for a simple but more effective tool to identify TB suspects as opposed to simple score or rule-based methods.

In developing countries, affording state-of-the-art technology such as using polymerase chain reaction (PCR) devices is still a hard thing to do. Nevertheless, these countries are the ones where the number of TB cases is still quite high. According to the Global Tuberculosis Report 2016 [5], “Six countries accounted for 60% of the new cases: India, Indonesia, China, Nigeria, Pakistan and South Africa.” Consequently, we argue that a lower-priced yet effective identification technique is necessary. This work addresses that particular challenge, where the best algorithm can then be installed on many devices, including smartphones, to help medical practitioners screen TB suspects.

This idea obviously comes with several challenges. For instance, most medical diagnosis cases have to deal with the imbalanced dataset problem. Since the majority of the people are healthy, the classification results are more likely to be negative. On the other hand, misclassifying people as TB positives (false positives) are considered less harmful than not being able to find the real positive cases (false negatives). Thus, our goal is to keep the sensitivity as high as possible while trying to increase the specificity.

We utilised several machine learning algorithms that are “most likely to be the bests” [6], such as support vector machines (SVM) with Gaussian (RBF) kernel. We also used XGBoost as one of the state-of-the-art algorithms for structured datasets and logistic regression for a simpler algorithm to compare. We evaluated our models by comparing our predictions with the corresponding GeneXpert®MTB/Rif results. Two score-based classification methods were used as the baselines in this study.

## II. Cohort

The dataset in this study was collected by several field workers. There are 37,856 data points collected from verbal screening. The data were collected from 12 November 2013 to 1 July 2016 from several sites in Jakarta, Indonesia.

### A. Cohort Selection

The people we screened came from both low-risk and high-risk settings. We used three strategies to collect our data. For the first strategy, we collected the data from people who were accompanying their relatives in hospitals or general practitioners, people in workplaces or industries, and inmates from several prisons in Jakarta.

Secondly, we also collected the data, without any screening process, from hospitals. These people were patients of the hospitals. We just asked them to provide their sputum samples to be tested as well.

For the third strategy, we cooperated with primary health centres, known as “puskesmas” in Indonesia, to retest some of the sputum samples. These samples are the ones which were already classified as TB-, using GeneXpert®MTB/Rif for confirmation. As it turns out, some of these people were incorrectly classified as TB-.

### B. Data Extraction

In the end, the dataset we are using in this experiment consists of 8732 rows with 7508 TB- and 1224 TB+ from the screening process with their available GeneXpert®MTB/Rif result. Most of them are not included since we did not test all of them for GeneXpert®MTB/Rif and did not go through verbal screening. We are also focusing on people with no history of TB prior to our study.

We can see later in this paper that some of the height data might be inaccurate. Hence, we chose to remove the data where the height is either unavailable or less than 120 cm. Even after we did this, we can still see that the height data are still doubtful.

We divided our dataset into three sets, i.e. training, validation, and test sets (60:20:20). The distribution of the classes for each dataset can be seen in Table I. We built our models using machine learning algorithms from the training set and then evaluated them with the validation set. Eventually, the best models from each algorithm were tested on the same test set to know which one performs the best in general.

**TABLE I.** Class distribution

|  | TB+ | TB- |
| --- | --- | --- |
| all | 1224 | 7508 |
| training set | 752 | 4487 |
| validation set | 234 | 1512 |
| test set | 238 | 1509 |

### C. Feature Choices

There are several attributes used in this experiment. These attributes are symptoms, comorbidities, and characteristics commonly found in TB patients. For example, we defined active smokers as people who have smoked more than 100 cigarettes in their lifetime.

Three attributes in this dataset are of numerical values, i.e. height, weight, and cough duration (in days). The height and weight were supposed to be measured by the screeners.

However, as depicted in Figure 1a, some of the height data might be estimated instead of actually measured. We can clearly see that rounded values like 160, 165, and 170 appear more often than the rest of the values.

**Fig. 1.**
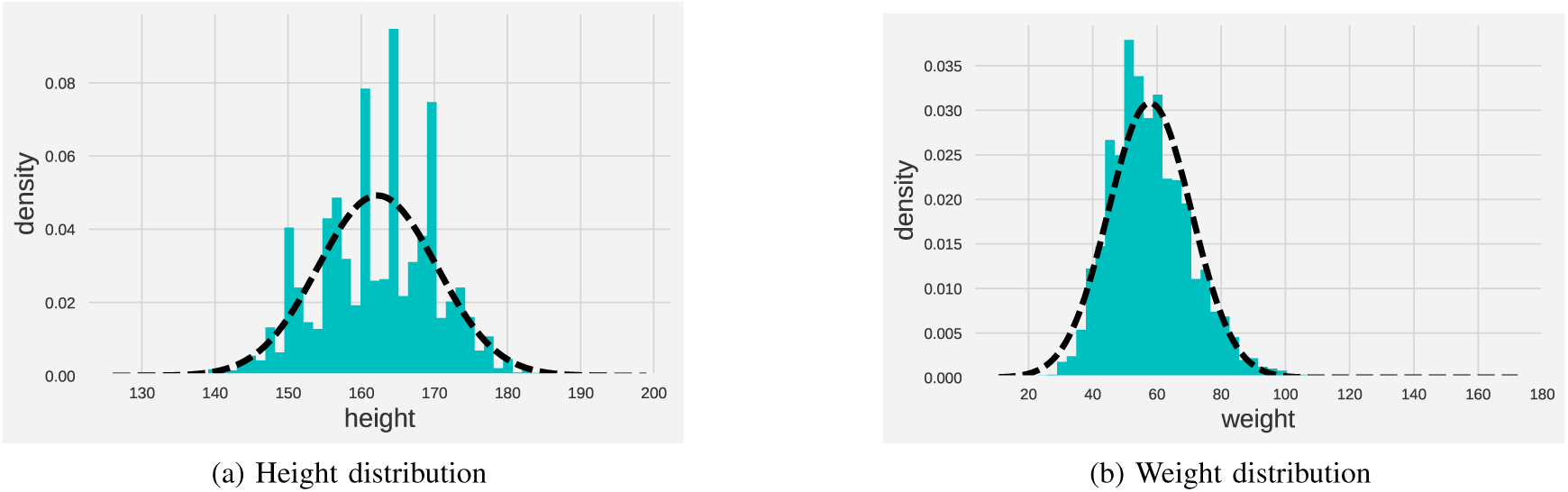
Measured attributes

On the other hand, the weight data appear to be more normally distributed. After we calculated the body mass index (BMI) from the height and the weight, we can see that the class-conditional Gaussians of the BMIs still correctly depict our expectation. In this case, we expect people with TB+ will be more likely to have lower BMI as one of their symptoms. Figure 2 confirms that we can still use the height and weight data to identify TB suspects. We also used the age and sex of the people screened as the attributes for machine learning algorithms.

**Fig. 2.**
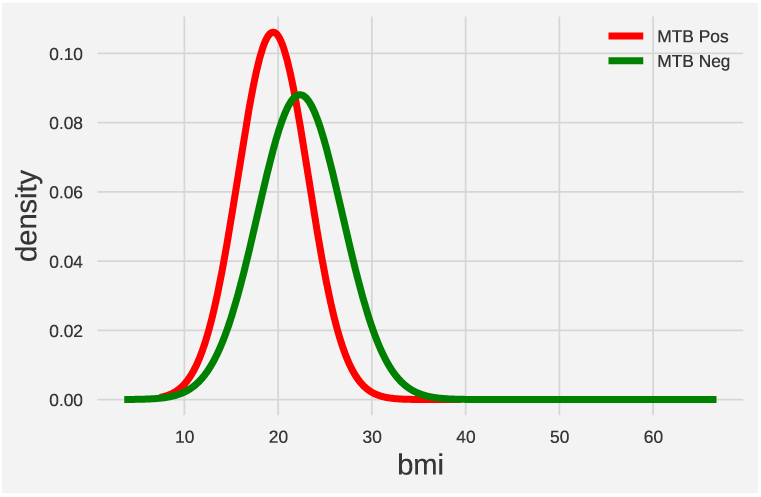
Class-conditional Gaussians from BMI

Aside from derived attributes like the body mass index (BMI), the attributes were acquired by verbally asking the subjects. We only marked ‘Yes’ as the value of diabetes, kidney failure, asthma, COPD, and HIV questions if the subjects had done the appropriate lab test before. The statistical properties of the dataset can be seen in Table II.

**TABLE II.** Statistical Properties of the Dataset

| Feature | Yes | No | Unknown |
| --- | --- | --- | --- |
| cough | 5480 | 3242 | 10 |
| fever | 1777 | 6955 | 0 |
| haemoptysis | 646 | 7758 | 0 |
| night sweat | 1365 | 7367 | 0 |
| weight loss | 1890 | 6842 | 0 |
| fatigue | 2185 | 6547 | 0 |
| breathing shortness | 189 | 1370 | 7173 |
| chest pain | 123 | 1436 | 7173 |
| diabetes | 969 | 5092 | 2671 |
| family diabetes | 137 | 468 | 8127 |
| TB exposure | 1414 | 4808 | 2510 |
| kidney failure | 75 | 4880 | 3777 |
| asthma | 717 | 4831 | 3184 |
| COPD | 232 | 4710 | 3790 |
| HIV | 251 | 4695 | 3786 |
| active smoker | 4313 | 4419 | 0 |

## III. Methods

### A. Score-based Classification

The clinicians who work with our company devised two scoring methods that were subsequently used throughout the project. When the total score of a person exceeds a certain threshold, we then classify that person as a TB suspect. Those TB suspects were then tested by GeneXpert®MTB/Rif to confirm our finding. The pseudo-code for these methods can be seen in Algorithm 1 and Algorithm 2 in Appendix. In these two algorithms, a person will be categorised as *underweight* if their BMI is lower than 18.5.

These two methods were designed to be loose, i.e. maximising the sensitivity of the classifier. However, as we later confirmed, the specificity could get quite low. In other words, this means that a lot of people got tested but turned out to be TB-. Moreover, Algorithm 2 was stricter and, in turn, produced lower sensitivity.

As mentioned earlier, this method needs experts to define the scoring scheme, i.e. how each attribute contributes to our suspicion about whether a person is TB+. Moreover, as the bacteria might mutate over time, the symptoms shown from a patient could be different from the time the scoring scheme is formulated. Thus, the scoring scheme would get outdated and should be redefined. To deal with this problem, we proposed evolving identification method by using machine learning classification which will learn as the number of data grows larger. In the next section, we describe some of the algorithms we used in this study.

### B. Machine Learning Classification

#### 1) Support Vector Machine

Support vector machines (SVM) try to linearly separate the data using a hyperplane. This can be done by finding the maximum margin solution with some help from kernel tricks. The maximum margin solution is defined as a constrained optimisation problem which can be solved by using Lagrange multipliers and quadratic programming. This will lead us to the weight vector **w** which is defined as

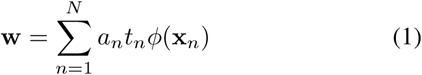

where *a*_*n*_’s are numerically determined non-negative coefficients and *t*_*n*_’s are the target values. In most cases, we will face the problem of non-linearly separable data. This is where *ϕ*(**x**_*n*_) as the feature space mapping of the input values will be useful [7].

The weight vector **w** along with the bias term *b* can then be used as the hyperplane to get the class of a new data point, which is formulated as

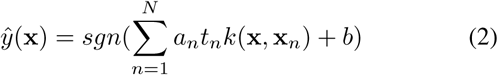

where *k*(**x, x**_*n*_) = *ϕ*(**x**) · *ϕ*(**x**_*n*_) is also known as the kernel function. There are many forms of kernel function, but in this study we are using the Gaussian or RBF kernel which can be formulated as in Eq. 3. As stated in [6], this particular kernel performs quite well on many cases.

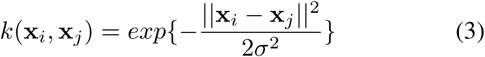

#### 2) XGBoost

One of the latest machine learning algorithms that is widely used nowadays is XGBoost [8]. This algorithm is a type of decision tree classifier that uses tree boosting method [9, pp. 337-388]. It also shares some characteristics with random forests, such as the ensemble trees concept.

It is also further stated in [8] that the “results demonstrate that our system gives state-of-the-art results on a wide range of problems.” One of the tasks where this algorithm works well is in structured datasets such as the one we are using in this study. Therefore, we will also use this in addition to the two aforementioned machine learning algorithms.

#### 3) Logistic regression

As a comparison, we also used a rather simple classifier in this study. We utilised a regularised logistic regression for binary class. We tried both L1 and L2 regularisation in the training step to determine the best outcome.

## IV. Results

### A. Evaluation Approach/Study Design

In this study, we compared two methods of classification, score-based methods as opposed to machine learning algorithms, to identify TB suspects from verbal screening data. We used sensitivity and specificity as the two main metrics to evaluate the algorithms. In medical screening cases, we prefer high sensitivity to high specificity due to the fact that we will confirm our screening results later on using lab tests. These metrics are formulated as,

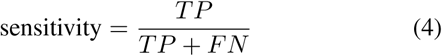

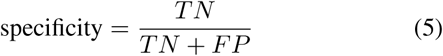

where *TP, TN, FP, FN* denote true positives, true negatives, false positives, and false negatives respectively.

Sensitivity is also known as true positive rate (TPR), whereas from specificity we can get false positive rate (FPR) which is equal to 1 − specificity. From these two metrics, we can then plot the receiver operating characteristic (ROC) curve to know which of the classifiers is generally better than the others. We can also compute the area under the curve (AUC) score from the ROC curve to get a single numerical value to compare the classifiers.

For the machine learning algorithms, we trained the untuned version of each classifier first to get an idea of how cost-sensitive approach would later improve our models. In the next step, we did the hyperparameter tuning with the parameters in Table III. After that, we trained the models using different weighting of positive class with the chosen hyperparameters.

**TABLE III.** List of hyperparameters

| Classifier | Hyperparameter | Values tested |
| --- | --- | --- |
| SVM | C | $[2^{-5}, 2^{-3}, 2^0, 2^3, 2^5, 2^7, 2^9]$ |
| | gamma | $[2^{-9}, 2^{-7}, 2^{-5}, 2^{-3}, 2^0, 2^3, 2^5, 2^7, 2^9]$ |
| XGBoost | max_depth | [3, 4, 5, 6, 7, 8, 9, 10] |
|  | min_child_weight | [0, 0.1, 1, 100, 1000] |
|  | gamma | [0, 0.1, 1, 100, 1000] |
| LogReg | penalty | ['l1', 'l2'] |
|  | C | [0.01, 0.1, 1, 10, 100] |
|  | max_iter | [100, 200, 300, 400, 500] |

### B. Preliminary Experiments and Hyperparameter Tuning

We first tried the aforementioned algorithms to our dataset. We kept the parameters to the default settings and imputed the unknown/null values to the most frequent values. Unsurprisingly, the classifiers tend to favour the negative class as can be seen in Table IV. The machine learning classifiers performed far worse than the score-based classifiers.

**TABLE IV.** Unweighted Classifiers Performance

| Classifier | Sensitivity (%) | Specificity (%) |
| --- | --- | --- |
| LogReg | 16.24 | 97.49 |
| XGBoost | 23.93 | 97.49 |
| SVM-RBF | 0.00 | 100.00 |
| Score-1 | 93.16 | 20.64 |
| Score-2 | 84.19 | 46.30 |

Aspiring to achieve a better result, we tuned the hyperparameters with several different class weights, such as 10, 20, 30, 40, and 50. We did grid search with 3-fold cross validation using our training data to get the optimal AUC score. AUC score was chosen because we want to optimise both sensitivity and specificity.

### C. Cost-sensitive Classification

We weighted the positive class to be 10, 15, 20, 25, 30, 35, 40, 45, and 50 when building the models. This turns out to be effective as shown in Figure 3. We even got a better sensitivity than the score-based classifiers while still maintaining the specificity score using this class weighting scheme.

**Fig. 3.**
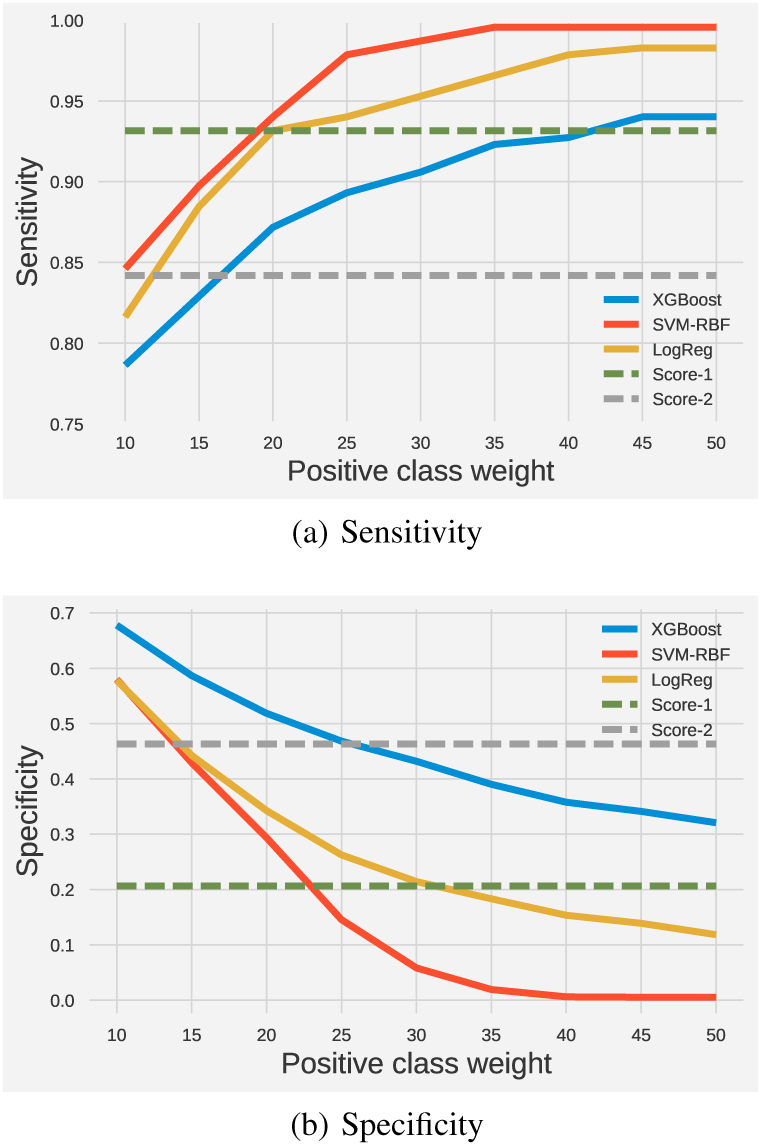
Performance of the classifiers

In SVM case, we got higher sensitivity than Score-2 and, simultaneously, higher specificity than Score-1 when we used at least 15 as the positive class weight. While the class weight is less than 20, we can also produce at least the same specificity as Score-1 with SVM. With XGBoost, we got an even better result: increasing the **specificity** from 20.63% to **35**.**78%** while only reducing the **sensitivity** from 93.16% to **92**.**74%** based on Score-1’s result (class weight: 40). Yet, this specificity is still 10.45% away from Score-2 specificity (46.23%). Logistic regression also produces a quite decent result when using 20 as the class weight, i.e. 93.16% sensitivity and 34.26% specificity. The ROC curve for the machine learning algorithms can be seen in Figure 4.

**Fig. 4.**
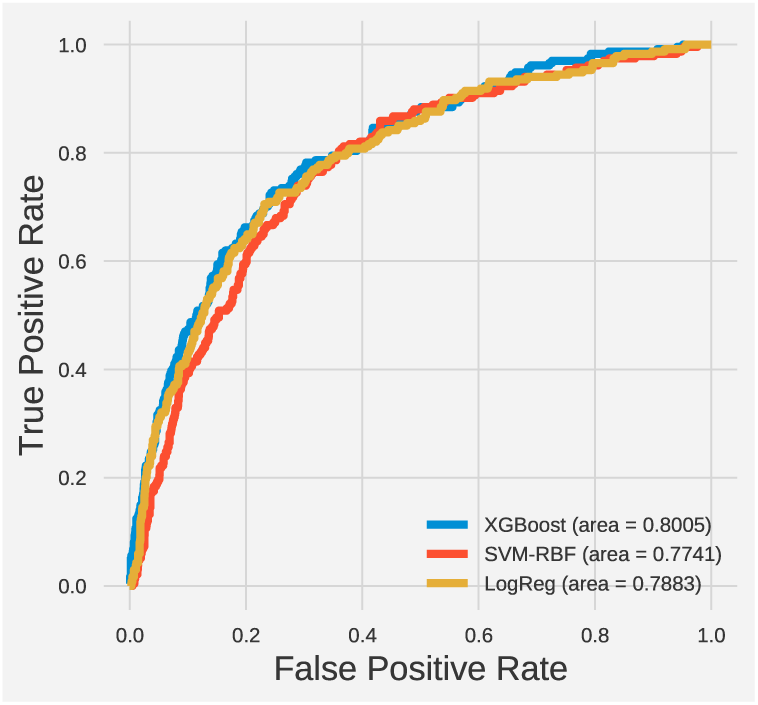
ROC curves

### D. The Effect of Dataset Size

One of the main advantages of machine learning algorithms over score-based methods is that we can hope for a better classifier as the number of data increases. We verified this assumption by increasing the number of training data gradually and tested the model to our validation set. In this experiment, we only used XGBoost as our classifier.

As shown in Figure 5, all the metrics seem to converge when the number of training data is ≈ 2000. This result implies that it might not be useful to keep feeding more data into the machine learning algorithms if we are looking for better classifiers. On the other hand, this also means that if we are trying to apply this method to different datasets, we might only need around 2000 data points to build a reasonably good model.

**Fig. 5.**
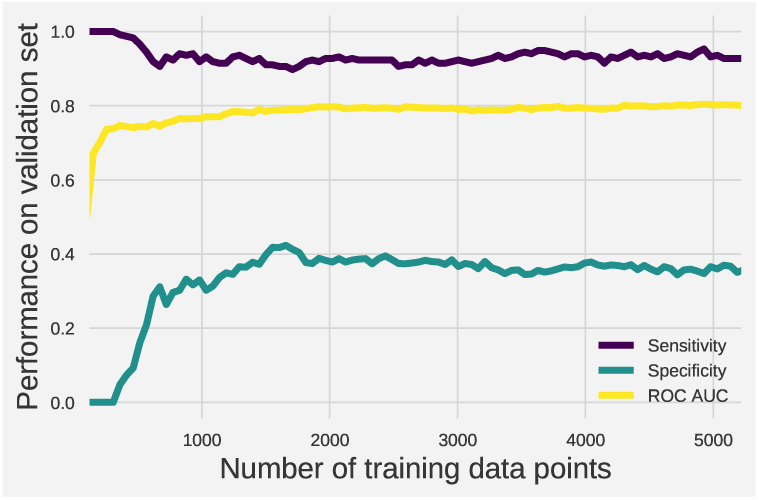
Dataset size effect on our chosen metrics

Our exploration with subsampling method also turned out to be ineffective. Since we are dealing with first-line patient recruitment through the screening process, we should seek for highly sensitive classifiers. However, the results from using subsampling method show otherwise, as can be seen in Table V. This shows that using cost-sensitive classifiers is preferred to using subsampling to overcome the imbalanced class problem.

**TABLE V.** Performance from Subsampling

| Classifier | Sensitivity (%) | Specificity (%) | ROC-AUC (%) |
| --- | --- | --- | --- |
| XGBoost | 68.80 | 74.27 | 78.39 |
| SVM | **73.50** | 71.03 | **78.63** |
| LogReg | 68.80 | **74.74** | 78.04 |

### E. Evaluation on Test Set

We took the best models from each algorithm and evaluated them on our test set. We also provided the results from the scoring methods. The performance of our models can be seen in Table VI.

**TABLE VI.** Performance on Test Set

| Classifier | Sensitivity (%) | Specificity (%) | Accuracy (%) |
| --- | --- | --- | --- |
| XGBoost | 96.64 | 35.06 | 43.45 |
| SVM | 96.64 | 29.82 | 38.92 |
| LogReg | **98.32** | 22.80 | 33.09 |
| Score-1 | 95.38 | 21.87 | 31.88 |
| Score-2 | 88.24 | **43.94** | **49.97** |

Although the best specificity and accuracy were achieved by the Score-2 method, the sensitivity is quite low. Recall that this is an early stage of TB diagnostics, thus our goal is to maximise the sensitivity first. Therefore, our best compromise is to use the XGBoost method where the sensitivity, specificity, and accuracy are the second best overall.

## V. Conclusions

As can be seen in the results, we were able to increase the specificity while still keeping the sensitivity relatively high by using machine learning algorithms. It also became apparent that using unbalanced weighting for the classes is necessary for this kind of task since we only got promising results after changing the class weights.

Building the inference models using these machine learning algorithms will need a certain number of data points before it stops improving in performance. In our case, the best algorithm converged after around 2000 data points. Therefore, we might use this as a reference point in further experiments. We might also need to incorporate other attributes, such as social-economy status, aspiring to get more interesting relationships in the future. This is because social-economy status can be retrieved from verbal screening and presumably independent from the current attributes we have used. A repository can then be built to store the different datasets and models for future work.

As it might suggest, increasing the specificity by more than 10% means saving lots of money and time to do unnecessary tests. This can be achieved without losing any sick people in the screening process. On the other hand, we do not really want maximum unconstrained sensitivity, since we can get this easily by classifying all the instances as TB+.

In a more practical sense, the best model from our work can be installed into smartphones where verbal screeners can rapidly identify TB suspects, especially in rural and poor areas where access to clinics might be limited. The suspects can then be taken to better health facilities to be tested further to confirm the findings. Although we could not achieve as good results as shown in [3], we have shown that we can also rely on data from verbal screening as well to get a reasonably well sensitivity. This method can be improved further when the medical records of a suspect can be used to answer the comorbidity questions.

## Data Availability

Not available for public

## APPENDIX

### Algorithm 1 Score-1

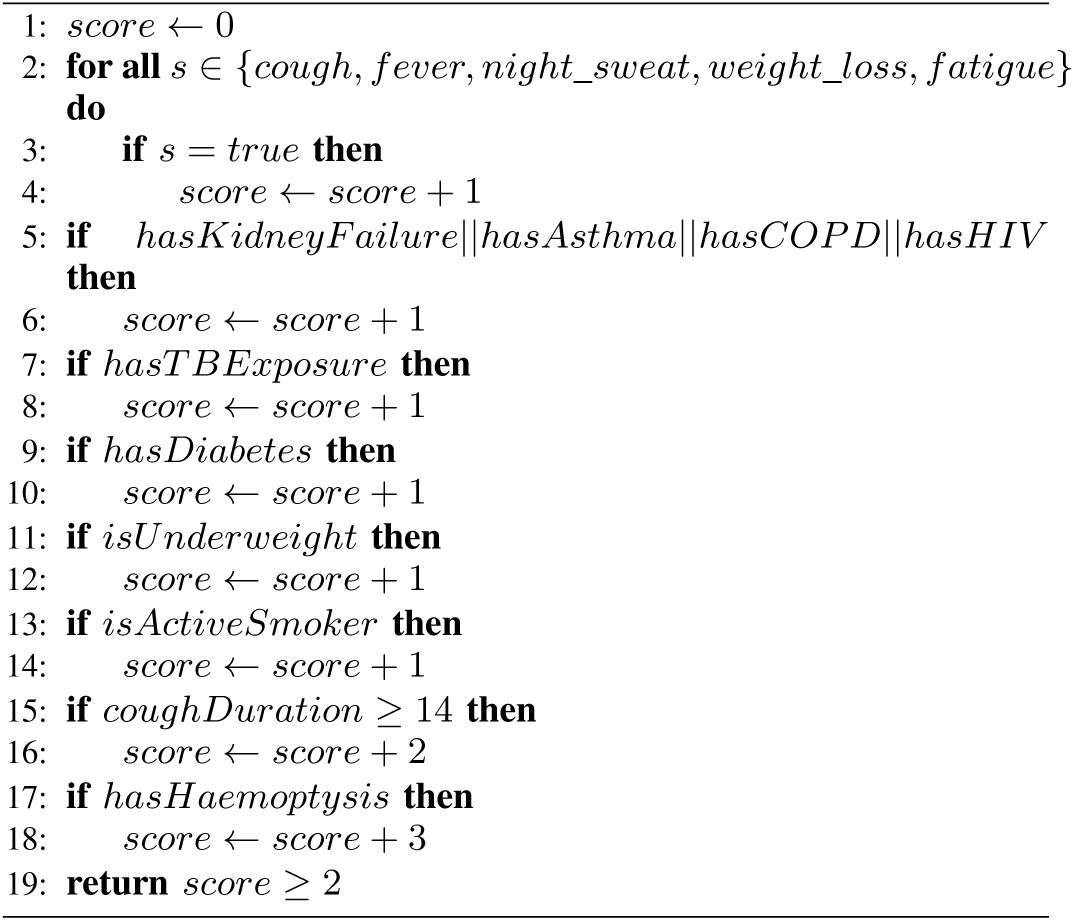

### Algorithm 2 Score-2

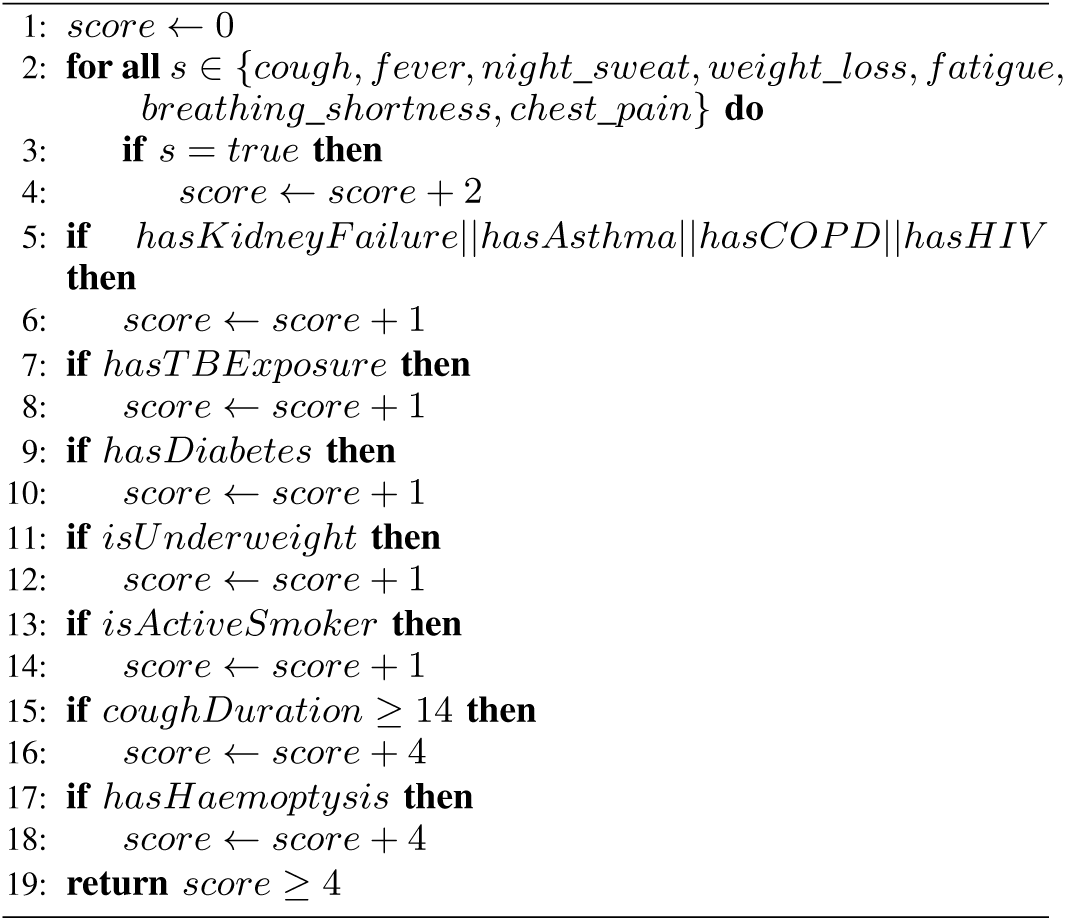

## References

[1] G. Hripcsak, C. A. Knirsch, N. L. Jain, and A. Pablos-Mendez, “Automated tuberculosis detection,” Journal of the American Medical Informatics Association, vol. 4, no. 5, p. 376, 1997. [Online]. Available:+http://dx.doi.org/10.1136/jamia.1997.0040376

[2] S. Shamshirband, S. Hessam, H. Javidnia, M. Amiribesheli, S. Vahdat, D. Petković, A. Gani, and M. L. M. Kiah, “Tuberculosis disease diagnosis using artificial immune recognition system,” Int J Med Sci, vol. 11, no. 5, pp. 508–14, 2014.

[3] J. Seixas, J. Faria, F. Souza, A. Vieira, A. Kritski, A. Trajman et al., “Artificial neural network models to support the diagnosis of pleural tuberculosis in adult patients,” The International Journal of Tuberculosis and Lung Disease, vol. 17, no. 5, pp. 682–686, 2013.

[4] C. Weng, C. Batres, T. Borda, N. G. Weiskopf, A. B. Wilcox, J. T. Bigger, and K. W. Davidson, “A real-time screening alert improves patient recruitment efficiency,” in AMIA Annual Symposium Proceedings, vol. 2011. American Medical Informatics Association, 2011, p. 1489.

[5] W. H. Organization et al., “Global tuberculosis report 2016,” 2016.

[6] M. Fernández-Delgado, E. Cernadas, S. Barro, and D. Amorim, “Do we need hundreds of classifiers to solve real world classification problems,” J. Mach. Learn. Res, vol. 15, no. 1, pp. 3133–3181, 2014.

[7] C. M. Bishop, Pattern Recognition and Machine Learning. New York, NY, USA: Springer, 2006.

[8] T. Chen and C. Guestrin, “Xgboost: A scalable tree boosting system,” in Proceedings of the 22nd ACM SIGKDD International Conference on Knowledge Discovery and Data Mining, ser. KDD ‘16. New York, NY, USA: ACM, 2016, pp. 785–794. [Online]. Available: http://doi.acm.org/10.1145/2939672.2939785

[9] T. Hastie, R. Tibshirani, and J. Friedman, The Elements of Statistical Learning, ser. Springer Series in Statistics. New York, NY: Springer New York, 2009. [Online]. Available: http://link.springer.com/10.1007/978-0-387-84858-7

